# Sleep spindle architecture associated with distinct clinical phenotypes in older adults at risk for dementia

**DOI:** 10.1101/2023.07.03.23292167

**Authors:** Isabella F. Orlando, Claire O’Callaghan, Aaron Lam, Andrew C. McKinnon, Joshua B. Tan, Johannes C. Michaelian, Shawn D.X. Kong, Angela L. D’Rozario, Sharon L. Naismith

## Abstract

Sleep spindles are a hallmark of non-REM sleep and play a fundamental role in memory consolidation. Alterations in these spindles are emerging as sensitive biomarkers for neurodegenerative diseases of ageing. Understanding the clinical presentations associated with spindle alterations may help to elucidate the functional role of these distinct electroencephalographic oscillations and the pathophysiology of sleep and neurodegenerative disorders. Here, we use a data-driven approach to examine the sleep, memory and default mode network connectivity phenotypes associated with sleep spindle architecture in older adults (mean age = 66 years). Participants were recruited from a specialist clinic for early diagnosis and intervention for cognitive decline, with a proportion showing mild cognitive deficits on neuropsychological testing. In a sample of 88 people who underwent memory assessment, overnight polysomnography and resting state fMRI, a k-means cluster analysis was applied to spindle measures of interest: fast spindle density, spindle duration and spindle amplitude. This resulted in three clusters, characterised by preserved spindle architecture with higher fast spindle density and longer spindle duration (Cluster 1), and alterations in spindle architecture (Clusters 2 and 3). These clusters were further characterised by reduced memory (Clusters 2 and 3) and nocturnal hypoxemia, associated with sleep apnea (Cluster 3). Resting state fMRI analysis confirmed that default mode connectivity was related to spindle architecture, although directionality of this relationship differed across the cluster groups. Together these results confirm a diversity in spindle architecture in older adults, associated with clinically meaningful phenotypes, including memory function and sleep apnea. They suggest that resting state default mode connectivity during the awake state can be associated with sleep spindle architecture, however this is highly dependent on clinical phenotype. Establishing relationships between clinical and neuroimaging features and sleep spindle alterations, will advance our understanding of the bidirectional relationships between sleep changes and neurodegenerative diseases of ageing.

## Introduction

Sleep spindles are a hallmark of non-rapid eye movement (NREM) sleep and are crucial for facilitating memory consolidation and learning (1–3). Alterations in these spindles are emerging as sensitive biomarkers for sleep disorders and neuropsychiatric conditions, including neurodegenerative diseases of ageing. In dementia and the precursor stages of mild cognitive impairment, alterations in sleep spindle architecture are observed (4–7). Identifying the clinical presentations associated with spindle alterations in older adults will be key to understanding the bidirectional relationships between sleep disturbance and cognitive disorders, and may inform early dementia diagnosis and selective interventions targeting sleep.

Sleep spindles evolve with a characteristic profile of transient sinusoidal cycles (typically 11 to 16 Hz lasting ∼0.5 to 3 seconds) on electroencephalography (EEG) during NREM sleep. Generated by thalamocortical circuits and regulated by the thalamic reticular nucleus (8,9), sleep spindles are nestled among the slow (low frequency power) waves that spread across the cortex during NREM sleep. Spindles modulate many aspects of thalamocortical communication, altering sensory transmission by inhibiting incoming sensory information to the thalamus (10–12), influencing cortical plasticity and synchronising hippocampal rhythms (i.e., sharp wave ripples) (13,14). Two types of sleep spindles, fast (>13 Hz) and slow (<13 Hz) (15), couple to different points throughout slow wave oscillations and are posited to have different functional roles (16). A growing body of evidence suggests that spindles, in particular fast spindles, undergo alterations in healthy ageing and are markedly decreased in Alzheimer’s disease and sleep disorders such as sleep apnea (6,17,18).

Early theories surrounding the functional significance of spindles centred on their role in inhibiting external sensory input during sleep (19,20), while more recent findings have identified their role in hippocampal-cortical memory processes (21,22). Memory consolidation is facilitated by the coupling of slow oscillations across the cortex, fast spindles, hippocampal sharp wave ripples and specific neuromodulatory states during NREM sleep (23–26). Interactions between the cortex and hippocampus are temporally tuned by slow oscillations and bursts of noradrenaline that synchronise hippocampal ripples with fast spindles (14,27–30). This evidence for sleep dependent memory consolidation supports the two stage model of memory, whereby memories are proposed to transfer from the hippocampus to the cortex for more permanent storage (2,31,32).

In this way, brain dynamics during sleep (i.e., fast spindles, hippocampal ripples, slow oscillations and locus coeruleus firing) may be viewed as markers of the memory consolidation process and its integrity. Relationships have been identified between these parameters, their coupling, memory, learning and disease. For example, higher fast spindle density is associated with consolidating certain types of memories (33), while the amplitude of spindles has been correlated with hippocampal activation after a visual learning task (34). Furthermore, ageing has been shown to impair the temporal coupling of slow oscillations and spindles, leading to impaired memory consolidation (35). These findings reaffirm the idea that spindles are fundamental mechanisms for reactivating and reprocessing recently acquired memory traces during sleep, via increasing hippocampal-cortical coupling. Across ageing, synchronisation of these NREM sleep rhythms changes considerably, with healthy older adults showing progressive reduction in spindle density, amplitude and duration (5). These changes are more prominent in mild cognitive impairment (18,36), Alzheimer’s disease (6,17,37) and Parkinson’s disease dementia (38). In individuals with mild cognitive impairment, shorter spindle duration and hippocampal atrophy are associated with reduced overnight memory consolidation (36).

Determining what drives changes in spindle architecture in neurodegenerative diseases compared to other conditions that affect sleep quality (e.g., sleep apnea), remains challenging – these conditions are known to co-occur with complex bidirectional relationships (39), with age being a major risk factor. For example, in ageing there is an increased prevalence of sleep apnea, a syndrome in which both spindle abnormalities and memory deficits can occur (40–42). In turn, poor sleep quality, as measured via self-report or objective polysomnography markers of sleep efficiency (e.g., wake after sleep onset, WASO), is observed in dementia (43) and preclinical Alzheimer’s disease (44), but it can also present in healthy ageing (45). Together, individuals with variable degrees of memory decline, sleep apnea, and reduced sleep quality may be key clinical phenotypes that can be associated with specific patterns of spindle alterations seen in ageing.

The extent to which changes in spindle architecture might relate to communication between brain regions involved in cognitive processes during the awake state is not fully understood. Simultaneous EEG functional Magnetic Resonance Imaging (fMRI) during sleep has shown that sleep spindles induce higher connectivity within thalamo-cortical-basal ganglia circuitry and the default mode network (46). Fast spindle density is related to enhanced hippocampal-cortical connectivity, as measured by fMRI the following day (47). The default mode network, which includes the hippocampal formation, lateral temporal cortex, posterior cingulate and inferior parietal lobules (48), is also a primary target for activity propagating from the hippocampus (49,50). Default mode network activation is linked with memory consolidation and recollective processes (51,52). Taken together, default mode network connectivity in ageing may be associated with sleep spindle architecture – offering insight into underlying interactions between sleep microarchitecture and large-scale network connectivity.

Here, we use a data driven approach to identify groups of older adults with distinct architecture in three sleep spindle parameters: fast spindle density, spindle duration and spindle amplitude. The sample comprised older adults attending a specialist clinic for early diagnosis and intervention for cognitive decline, with a proportion of them showing mild cognitive deficits on neuropsychological testing. To identify the clinical phenotypes associated with these groups, we characterised them with respect to key measures relevant in ageing: memory function, oxygen desaturation events (a measure of nocturnal hypoxemia and marker of sleep apnea severity) and sleep quality (WASO). We further explored associations between default mode network connectivity and sleep spindle parameters.

## Methods

### Participants

Participants were recruited from the Healthy Brain Ageing Clinic, University of Sydney, a specialist early intervention memory and cognition clinic for older adults. Inclusion criteria for the clinic were age ≥ 50 years and a general practitioner or specialist referral. Exclusion criteria were: history of psychiatric or neurological disorders; loss of consciousness >30 min; Mini-Mental State Examination score (MMSE; (53)) <24; history of drug and/or alcohol misuse; limited English proficiency; transmeridian travel within a week prior to the sleep study. The current study sample included all individuals attending between 2011-2020, who did not meet criteria for dementia (based on clinical and neuropsychological assessment), and who underwent both overnight polysomnography (PSG) and resting-state fMRI within 6 months. This resulted in a sample of 127 participants, then following exclusion of people with missing or corrupted PSG data, the final study sample was 88. The study was approved by the local ethics committee and all participants provided written informed consent.

### Clinical and memory assessment

Participants underwent full clinical, neuropsychological and mood assessment using a standardised assessment battery, described elsewhere (54). Medical history was obtained by a medical specialist, which included ruling out major sleep disorders. Body mass index (BMI; kg/m) and weekly alcohol consumption (standard drinks per week) were calculated. Psychiatric history was assessed via structured clinical interview questions (55), and participants self-reported depression symptoms on the Geriatric Depression Scale short form-15 item (56). Self-reported sleep quality was assessed using the Pittsburgh Sleepiness Quality Index (PSQI; (57)).

Memory performance, the focus of the current study, was assessed via: i) the Rey Auditory Verbal Learning Test (RAVLT; (58)), a 15-item list learning task with five learning trials followed by a 20 minute delay; ii) the Logical Memory subtest of the Wechsler Memory Scale III (59), a story learning task involving recall after a 25-35 minute delay. For both measures, delayed recall scores were used as the outcome of interest; these were z-scored and averaged to create a memory composite score. Learning and executive function performance are reported in the Supplementary Material for descriptive purposes. Premorbid IQ was derived from the Wechsler Test of Adult Reading (60).

Based on standardised criteria (61) and a consensus meeting of a medical specialist and two neuropsychologists, participants were classified as displaying mild cognitive deficits based on neuropsychological test performances >1.5 standard deviations below the age adjusted normative data and relative to premorbid IQ. Performance was further classified into four subtypes: 1) amnestic, single domain; 2) amnestic, multiple domain; 3) non-amnestic, single domain; 4) non-amnestic, multiple domain.

### Polysomnography and EEG analysis

During overnight polysomnography, EEG signals were recorded from Fz, F3, F4, Cz, C3, C4, Pz, O1, and O2 electode sites with contralateral reference to mastoids (i.e., F3 to M2, F4 to M1 and midline channels with reference to the average of M1 and M2) at sampling rates of 200 Hz (Alice 5, Philips, Netherlands) or 512 Hz (Embla Titanium, Natus, USA). Trained sleep technicians manually identified sleep stages and respiratory events according to standardised AASM 2.2 criteria (62).

Our spindle analysis focused on central EEG sites: C4-M1 was preferentially used and where C4-M1 data was missing or was poor quality, C3-M2 was used. Recordings underwent automated EEG artifact detection using a validated algorithm (63). Following artifact detection, an automated algorithm identified sleep spindle events with a duration of ≥ 0.5 ≤ 3 s with a mean frequency of 11–16 Hz in NREM sleep (N2 and N3). Validation of the automated spindle detection algorithm is previously reported (36). Fast spindle density (number of fast sleep spindle events (> 13 ≤ 16 Hz) per minute of NREM sleep), overall spindle duration (average duration in seconds of all sleep spindle events within the 11-16 Hz range) and overall spindle amplitude (average maximum amplitude µV of all sleep spindle events within the 11-16 Hz range) were the *a priori* spindle measures of interest. To characterise sleep quality we calculated wake after sleep onset (WASO) as the amount of time spent awake after sleep onset. For nocturnal hypoxemia as a marker of sleep apnea severity we calculated the oxygen desaturation index (ODI) based on the number of dips in oxygen saturation levels of 3% or more per hour of sleep. Other measures of sleep macroarchitecture (including sleep stage durations, sleep onset latency and sleep efficiency) are presented in Supplementary Material.

### K-means clustering

K-means clustering was performed on the three sleep spindle measures of interest (i.e., spindle duration, spindle amplitude and fast spindle density) using the *kmeans* function in R. To determine the optimal number of clusters we used a consensus-based algorithm, implemented using *n_clusters* from the ‘parameters’ R package (64). This algorithm compares several common approaches and finds the number of clusters most agreed upon across the different methods. To assess cluster stability, we ran a bootstrap stability measure with 50 iterations using the ‘bootcluster’ package in R (65). We calculated the average outputted Jaccard coefficient, a statistic used to assess the similarity of two clustering assignments, ranging from 0 (no overlap) to 1 (perfect overlap) (66). We ran this bootstrap test 100 times to obtain mean and confidence interval measures across all repetitions.

### Statistical analysis

Statistical analyses were conducted in R (version 4.2.1, (67)). Demographics and clinical screen scores were compared across the three cluster groups using chi-squared tests and one-way analysis of variance (with post hoc Tukey’s pairwise comparisons). We hypothesised that the clusters based on spindle architecture would relate to variables of interest: memory performance (memory composite score), sleep quality (WASO) and oxygen desaturation (3% ODI). We therefore conducted a multinomial regression in the ‘nnet’ package (68), using those variables of interest to explain group cluster membership. Multinomial logistic regression models estimate the probability of belonging to a particular group (in our case, belonging to a particular cluster) based on a set of explanatory variables. Age and sex were included as fixed effects in the model, and continuous variables were z-scored. Logistic regression coefficients are presented as β estimates, as well as odds ratios and 95% confidence intervals. *P*-values were calculated using Wald tests, that is, z-statistics were created using the ratios of model coefficients and standard errors, from which *p*-values were created using the standard normal distribution. Cluster 1 was set as the baseline/reference level. Negative β estimates suggest that as a variable increases, it is more likely to be predictive of the reference group (i.e., Cluster 1); whereas positive β estimates suggest that as a variable increases, it is more likely to be predictive of the comparison group (i.e., Clusters 2 or 3). An odds ratio of <1 further indicates that the probability of remaining in the comparison group falls with every unit increase of the variable; an odds ratio >1 indicates the probability of remaining in the comparison group becomes more likely with every unit increase of the variable.

### Neuroimaging analysis

#### Image acquisition

T1-weighted structural and resting state scans were obtained using a GE MR750 3-Tesla MRI (General Electric, USA). Structural MRI was acquired using used an 8-channel head coil, and a 3D-T1-weighted BRAVO Spoiled Gradient-Recalled sequence with 196 sagittal slices (repetition time = 7.2 ms; echo time = 2.8 ms; flip angle = 12°; matrix 256 × 256; 0.9 mm isotropic voxels). Resting-state fMRI was acquired using a T2*-weighted echo planer imaging sequence (39 axial slices covering the whole brain; TR = 3000 ms, TE = 36 ms, flip angle = 90°, matrix: 64 × 64, in-plane voxel size = 3.75 mm × 3.75 mm × 3 mm). The first five volumes were discarded to eliminate spurious T2-equilibration effects, before a further 140 echo planer imaging-volumes were acquired in a single run with eyes closed.

#### Resting state fMRI pre-processing and analysis

Pre-processing was performed in MATLAB using the functional connectivity toolbox (CONN v19.c; http://www.nitrc.org/projects/conn) which involved standard preprocessing steps including: co-registration, normalisation, unwarping, noise component extraction, segmentation, skullstripping and denoising. See Supplementary Material for a full description.

Following pre-processing, we extracted BOLD time series from 333 cortical regions (69) for 140 time points. A functional connectivity matrix was calculated for each individual using Pearson correlation values, producing a 333 × 333 matrix representing connectivity between regions. Each region was assigned to 12 canonical functional networks (69), and within network connectivity was calculated as the average connectivity across regions in a network.

To identify whether default mode network connectivity was associated with spindle architecture, we constructed linear models in R and used the ‘emmeans’ package (70) for *post hoc* comparisons including Tukey adjustments for multiple testing. In three separate linear models, we examined whether spindle architecture was related to default mode network connectivity, and whether this differed based on cluster membership; age and sex were included as fixed effects. An illustrative model setup in R syntax for spindle duration: `spindle duration ∼ DMN connectivity * cluster + age + sex`.

### Code availability

Code to reproduce manuscript figures and statistical analyses are freely available through the Open Science Framework (https://osf.io/3gqb9/).

## Results

### Overall sample characteristics

The final study sample consisted of 88 older adults (30 males/58 females), mean age 66 years (SD = 8.34, range = 50-86) and average 13 years of education (SD = 2.86, range = 7-9). Overall, 64 of the 88 participants displayed mild cognitive deficits on neuropsychological testing (3 = amnestic, single domain; 23 = amnestic, multiple domain; 19 = nonamnestic, single domain; 19 = nonamnestic, multiple domain).

### K-means clustering solution

An optimal solution of three clusters was identified by the consensus-based algorithm (see Supplementary Figure 1). The overall stability of this cluster scheme was 0.78 (95% CI = 0.785, 0.775), as validated under bootstrap replications. Results of the three cluster solution are shown in Figure 1.

**Figure 1.**
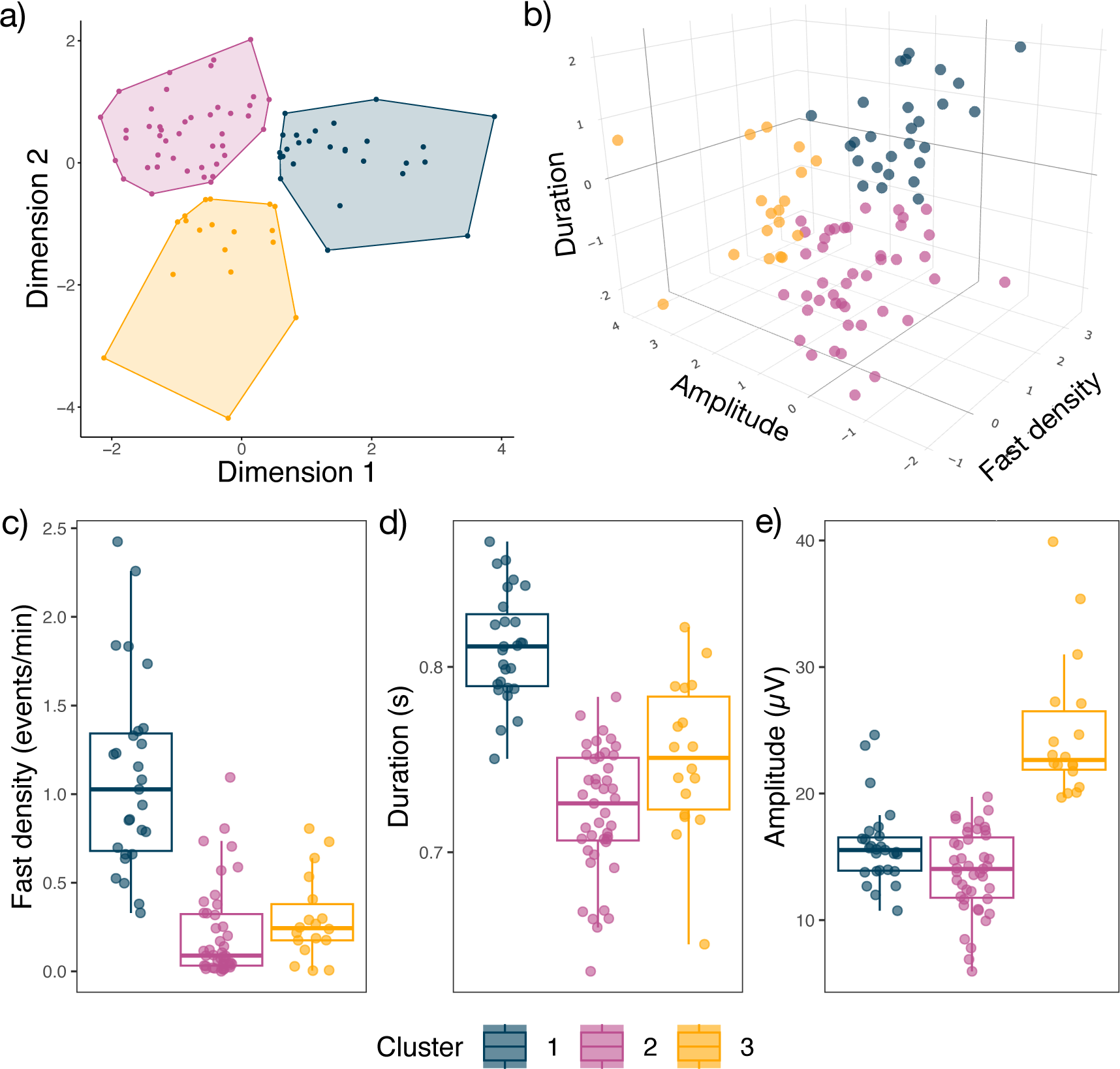
Clustering solution using sleep spindle parameters of fast spindle density, spindle duration and spindle amplitude. **a)** 2D visualisation of the three clusters (NB. a principal component analysis is performed to represent the variables in the 2D plane); **b)** 3D visualisation of cluster characteristics (interactive 3D plot can be explored here https://osf.io/69qa8); **c-e)** Cluster spindle characteristics, visualising the three variables making up each cluster.

Cluster 1 (n = 27) was characterised by a higher density of fast spindles (mean = 1.10, SD = 0.55 events per minute of NREM sleep) with longer spindle durations (mean = 0.81, SD = 0.03 seconds). Cluster 2 (n = 43) had the lowest density of fast spindles (mean = 0.21, SD = 0.26) and shortest spindle durations (mean = 0.72, SD = 0.03). Cluster 3 (n = 18) also had a low density of fast spindles (mean = 0.30, SD = 0.24) and shorter spindle durations (mean = 0.75, SD = 0.04), relative to Cluster 1 and had the highest spindle amplitudes (mean = 24.81, SD = 5.54, compared to Cluster 1; mean = 15.87, SD = 3.15, and Cluster 2; mean = 13.84, SD = 3.25 µV).

### Cluster demographic and clinical characteristics

Demographics and clinical screening measures are described in Table 1. Participants comprising the three clusters did not differ significantly in terms of sex, education, Geriatric Depression Scale score, self-reported sleep quality, BMI or alcohol consumption (*p* values > 0.07). Age, MMSE and the proportion of participants with mild cognitive deficits differed between clusters. For age [*F*(2, 85) = 4.5, *p* = 0.014], *post hoc* tests confirmed significantly older participants in Cluster 2 vs. 3 (*p*_adjusted_ = 0.033). MMSE also differed across the groups [*F*(2, 83) = 3.53, *p* = 0.034], *post hoc* tests confirmed significantly lower scores for participants in Cluster 2 vs. 1 (*p*_adjusted_ = 0.029). The proportion of members with mild cognitive deficits across the clusters differed significantly (*X*^2^ (2) = 7.82, *p* = 0.020). Cluster 2 had a higher proportion of people with mild cognitive decline compared to Cluster 1 (*p* = 0.052) and significantly higher proportion relative to Cluster 3 (*p* = 0.025). Variables of interest used in the subsequent multinomial regression models also showed significant differences between clusters. For the memory composite score [*F*(2, 85) = 9.49, *p* < 0.001], *post hoc* tests confirmed significantly lower scores for participants in Clusters 2 (*p*_adjusted_ < 0.001) and 3 (*p*_adjusted_ = 0.008) relative to Cluster 1. Sleep quality (measured by WASO) differed across the groups [*F*(2, 85) = 4.63, *p* = 0.012], *post hoc* tests confirmed significantly poorer scores for participants in Cluster 2 (*p*_adjusted_ = 0.015) relative to Cluster 3. Oxygen desaturation index differed across the groups [*F*(2, 81) = 4.11, *p* = 0.020], *post hoc* tests confirmed significantly higher scores for participants in Cluster 3 relative to Clusters 1 (*p*_adjusted_ = 0.027) and 2 (*p*_adjusted_ = 0.031). Additional neuropsychological and sleep macroarchitecture characteristics for the clusters are described in Supplementary Material.

**Table 1.**
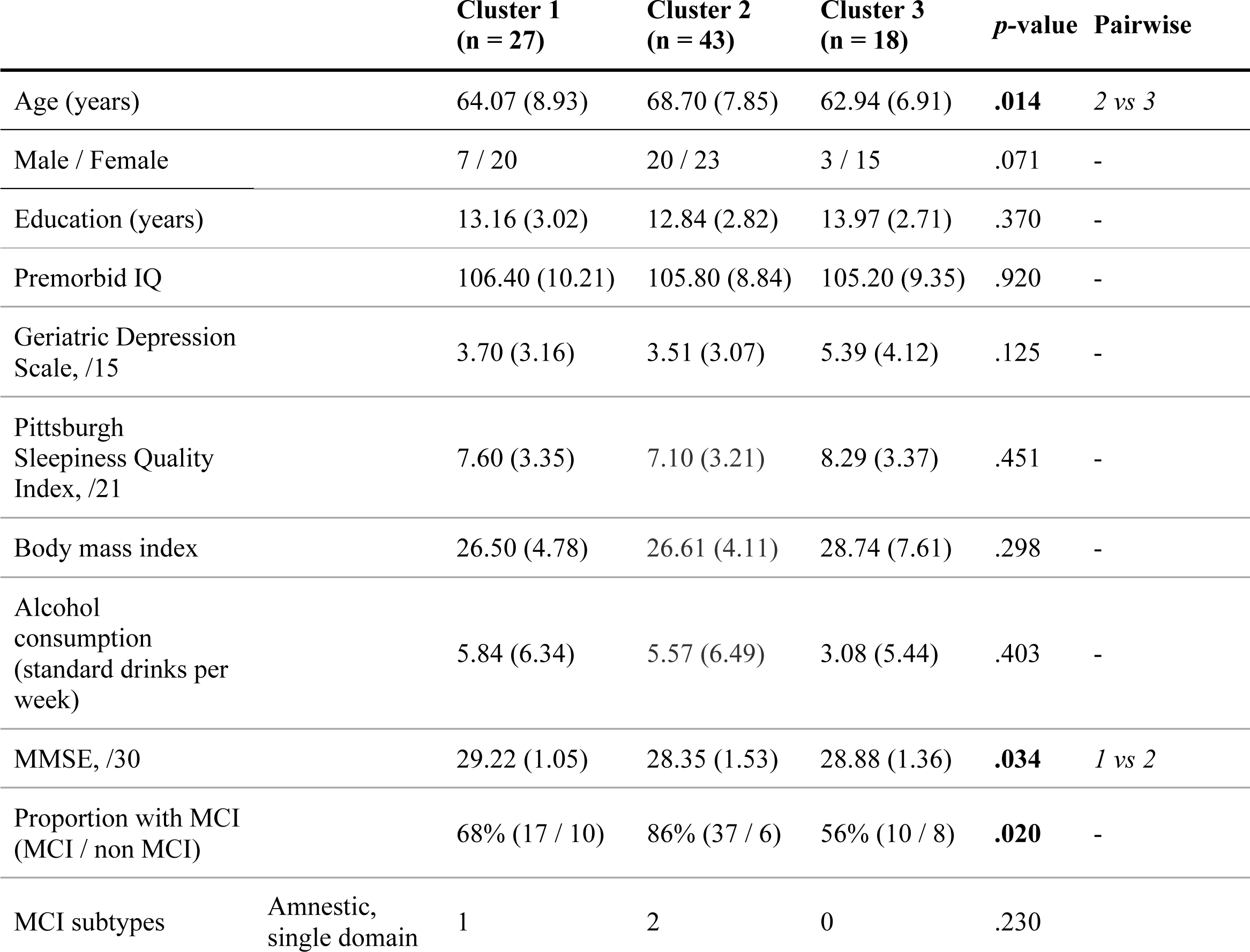
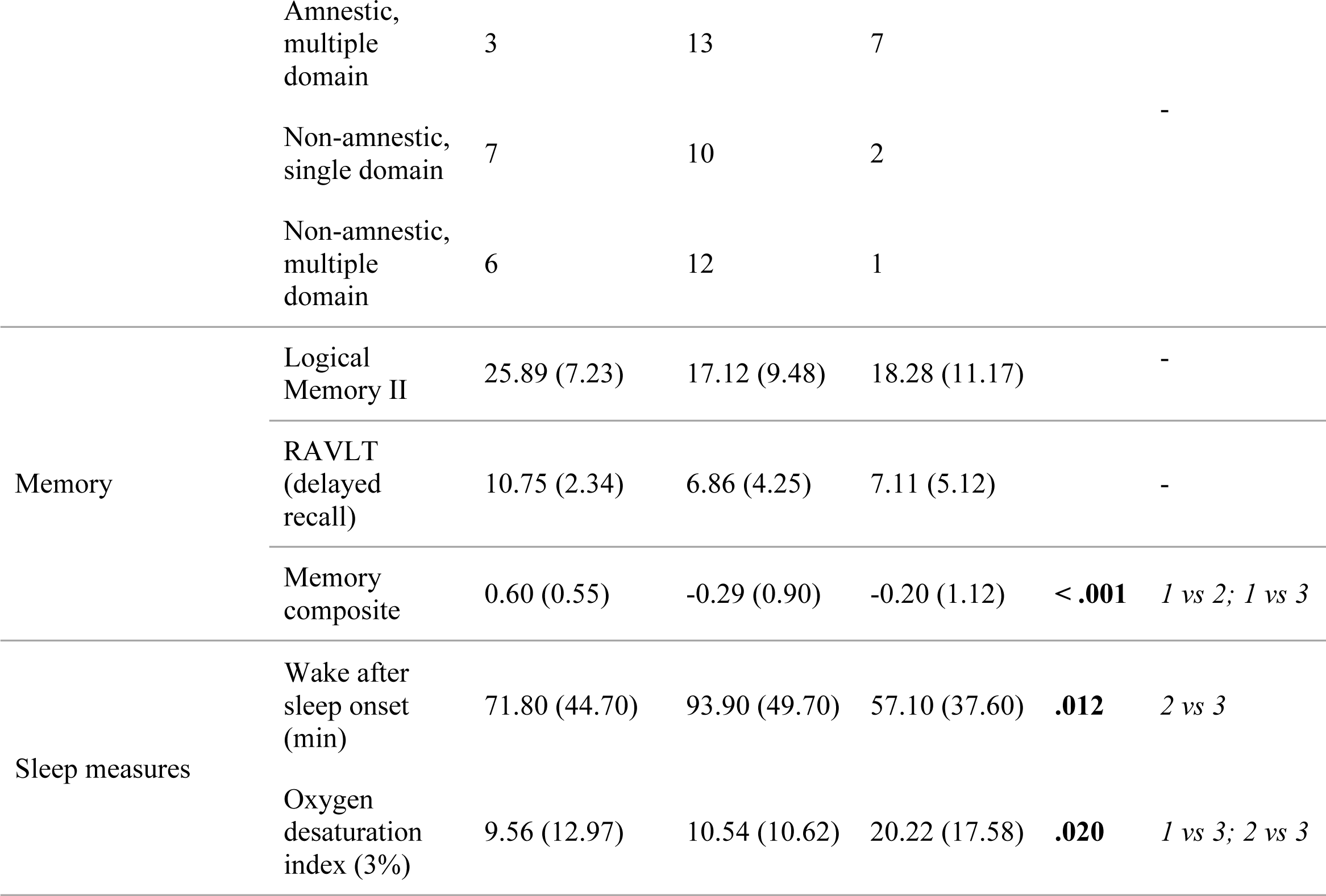
Characteristics of k-means defined cluster groups. Data are presented as mean (standard deviation). Group differences are compared using one-way analysis of variance with post hoc Tukey’s honestly significant difference or chi-squared tests. P-values shown for group level comparisons (significant post hoc results reported in the manuscript). MMSE = Mini-Mental State Examination; RAVLT = Rey Auditory Verbal Learning Test. Memory composite = average of z-scored Logical Memory II and RAVLT scores.

### Probability of cluster membership based on memory, sleep quality and apnea profiles

Results of the multinomial regression are shown in Table 2 and Figure 2. Cluster 1 (characterised by a higher density of fast spindles and longer spindle durations) was set as the baseline/reference level. Memory score emerged as a significant predictor of cluster membership. With every 1 unit increase in memory performance, the odds of being grouped in Cluster 2 decreased by 70% (OR: 0.30, 95% CI: 0.13-0.68) and of being in Cluster 3 by 82% (OR: 0.18, 95% CI: 0.06-0.51), after adjusting for other variables of interest, plus age and sex. Oxygen desaturation index was a significant predictor of Cluster 3 membership, such that every 1 unit increase was associated with a 7% increased likelihood of being grouped in Cluster 3 (OR: 1.07, 95% CI: 1.01-1.13), adjusting for sleep quality, memory score, age and sex. Sleep quality (measured by WASO) was not a significant predictor cluster membership (Cluster 2: OR = 1.01, 95% CI: 1.00-1.02; Cluster 3: OR = 0.99, 95% CI: 0.96-1.01).

**Figure 2.**
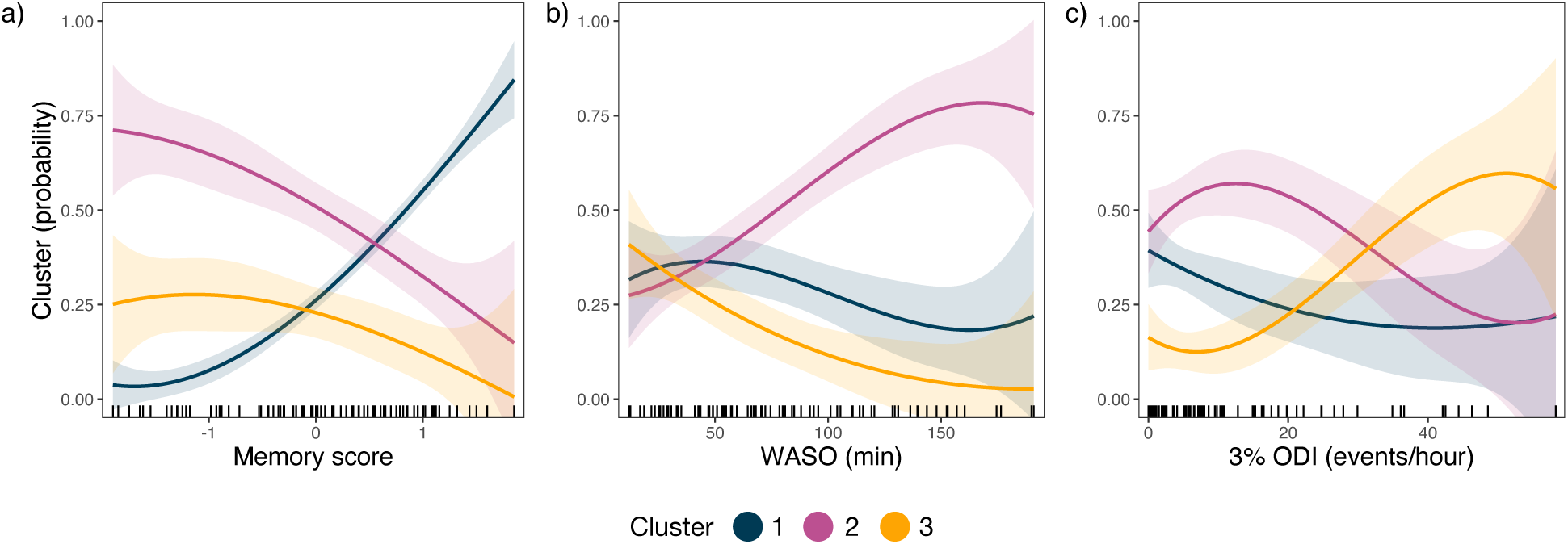
Cluster wise relationships with key clinical measures. Probability that an individual is in a given cluster (y-axis) along with their score on the variables of interest (x-axis). Ribbons show standard error of the probability estimate. **a)** Memory = composite memory score. **b)** WASO = wake after sleep onset (min). **c)** 3% ODI = 3% oxygen desaturation index (events per hour).

**Table 2.** Results of multinomial regression model, with age and sex included as fixed effects. Memory refers to the composite score. Sleep quality measured by WASO. CI = confidence interval. Oxygen desaturation index measured by 3% ODI events per hour.

| | | $\beta$ | Odds ratio | 95% CI | *p*-value |
| --- | --- | --- | --- | --- | --- |
| **Cluster 2\*** | Memory | -1.22 | 0.30 | 0.13-0.68 | **0.004** |
|  | Sleep quality | 0.01 | 1.01 | 1.00-1.02 | 0.157 |
|  | Oxygen desaturation index | -0.01 | 1.00 | 0.95-1.05 | 0.839 |
| **Cluster 3\*** | Memory | -1.74 | 0.18 | 0.06-0.51 | **0.001** |
|  | Sleep quality | -0.02 | 0.99 | 0.96-1.01 | 0.224 |
|  | Oxygen desaturation index | 0.07 | 1.07 | 1.01-1.13 | **0.018** |

\*Compared to Cluster 1 as reference level

### Default mode network connectivity associated with sleep spindle architecture and clinical phenotype

Linear models examined the relationship between default mode network connectivity and cluster membership on sleep spindle architecture, controlling for age and sex. For fast spindle density, there was a significant main effect of default mode network connectivity (F(1, 76) = 13.55, *p <* 0.001) and a significant interaction with cluster membership (F(2, 76) = 4.17, *p =* 0.019). As shown in Figure 3a, fast spindle density had distinct relationships to default mode network connectivity across the clusters. *Post hoc* tests confirmed that Cluster 1 significantly differed to Cluster 2 (β = −0.435, *p* = 0.018) but not to Cluster 3 (β = −0.332, *p* = 0.189) (Figure 3a). Clusters 2 and 3 did not differ significantly (β = 0.103, *p* = 0.854). For spindle duration, we also found a significant main effect of default mode network connectivity (F(1, 76) = 6.61, *p* = 0.012) and a significant interaction (F(2, 76) = 5.06, *p =* 0.008), suggesting spindle duration also has distinct relationships to connectivity depending on cluster membership (Figure 3b). *Post hoc* tests confirmed that Cluster 1 differed significantly to both Clusters 2 (β = −0.409, *p* = 0.032) and 3 (β = −0.516, *p* = 0.023), while Clusters 2 and 3 did not differ (β = −0.108, *p* = 0. 847) (Figure 3b). Together, these results indicate a pattern in Cluster 1 (i.e., increasing default mode connectivity associated with fewer faster spindles and shorter spindle durations), which is not present, or is indeed the opposite, in Clusters 2 and 3. These results were not evident for spindle amplitude, where there was no significant main effect of default mode network connectivity (F(1, 76) = 0.24, *p* = 0.626), nor was there a significant interaction with cluster membership (F(2, 76) = 0.33, *p =* 0.719).

**Figure 3.**
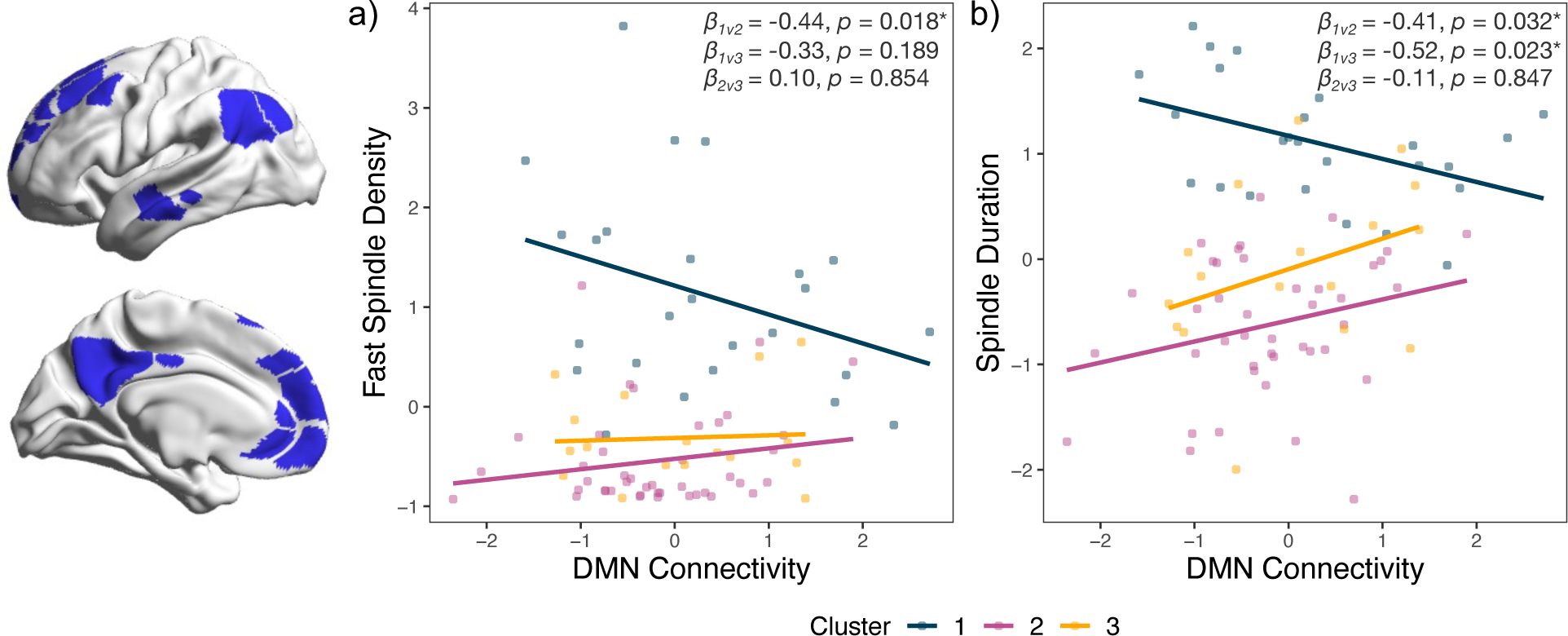
Default mode network connectivity associated with spindle architecture. **a)** Relationship of average default mode network (DMN) connectivity and fast spindle density (events per min during NREM sleep) for each individual, coloured by cluster membership. **b)** Relationship of average default mode network connectivity and spindle duration (sec) for each individual, coloured by cluster membership. Beta coefficients and p-values of the interaction effect for Clusters 2 and 3 against Cluster 1 are included in figures.

Given the distinct patterns that emerged across clusters for the default mode connectivity – spindle relationships, we confirmed whether these were unique to the default mode network. In this exploratory follow-up analysis, we conducted the same linear models for other networks associated with memory and cognition (i.e., the ventral attention, dorsal attention, frontoparietal, retrosplenial and cingulo opercular networks). No significant main effects or interaction effects with cluster membership emerged for sleep spindle fast density and duration (p > 0.05). Details are reported in the Supplementary material.

## Discussion

In a sample of older adults, we identified distinct clusters characterised based on features of sleep spindle duration, fast spindle density and spindle amplitude. The clusters were associated with unique clinical phenotypes relating to memory function and oxygen desaturation index. Furthermore, we showed that the relationships between spindle parameters and default mode network connectivity differed depending on cluster membership. Together, these findings suggest that distinct clinical phenotypes and neuroimaging signatures reflect underlying changes in sleep microarchitecture which may inform early identification of cognitive and neurodegenerative changes in ageing.

Of the three groups that emerged, Cluster 1 was characterised by spindles of the longest duration, highest fast spindle density and moderate amplitude. Cluster 2 was characterised by the shortest spindle durations, lowest density of fast spindles and lowest amplitude. Finally, Cluster 3 showed reduced density of fast spindles and shorter spindle durations, relative to Cluster 1, and the highest spindle amplitude. Multinomial regression analysis revealed that memory function and nocturnal hypoxemia due to sleep apnea (oxygen desaturation index) differed across the clusters. With respect to Cluster 1, Cluster 2 was significantly associated with reduced memory performance: people with the lowest memory scores had the highest probability to be in this cluster (Figure 2a). People in Cluster 2 were more likely to have poorer sleep quality as measured with higher wake after sleep onset (Figure 2b), though this did not emerge as a significant predictor. Cluster 3 was significantly associated with both reduced memory scores and higher oxygen desaturation index, with respect to Cluster 1. Age and sex were controlled for in these models, which is noteworthy given the significant age difference between Clusters 2 and 3.

Taken together, Cluster 1 emerged as a cohort with relatively preserved memory function, the longest spindle duration and highest fast spindle density. Cluster 2 was characterised by reduced memory performance, the highest proportion of people with mild cognitive deficits based on full neuropsychological testing, and the highest wake after sleep onset (suggesting poorer sleep quality), in the context of having the shortest spindle durations, lowest density of fast spindles and lowest spindle amplitude. Finally, Cluster 3 was characterised by reduced memory function and greater night-time hypoxemia (indicative of worse sleep apnoea severity), along with reduced fast spindle density, shorter spindle durations and the highest spindle amplitude.

Memory performance emerged as a key distinguishing feature of the groups. Sleep spindles are known to contribute to memory consolidation and retrieval (23), and as such may be a promising biomarker to supplement early dementia diagnosis and monitoring. Reduced fast spindle density and shorter spindle duration, exceeding that seen in ageing, have been found in mild cognitive impairment, Alzheimer’s disease and Parkinson’s disease dementia (6,17,18,36,38,71,72). Worsening global cognition has also been linked with reduced spindle duration (72), and a reduction in spindle density has been correlated with cerebrospinal fluid levels of amyloid-beta 42 (Aβ42), phosphorylated tau (p-Tau), and total-tau (T-Tau) markers of Alzheimer’s disease pathology (73). These findings suggest that sleep spindles are impaired in neurodegenerative diseases of ageing, including in the prodromal stages, and their architecture, namely fast spindle density and spindle duration, are associated with neurodegeneration markers and cognitive decline. Our Cluster 2 profile with the most reduced memory scores and altered spindle architecture (relative to Cluster 1), may identify individuals at greatest risk for dementia. Indeed, this group had a higher proportion of people with mild deficits detected on neuropsychological testing. Although it should be noted that our data is not equipped to discriminate between mild cognitive impairment due to Alzheimer’s disease/neurodegenerative disease versus stable mild cognitive impairment, which can be substantiated by CSF Aβ42, p-tau, and T-tau biomarkers (74,75).

While reductions in spindle duration, fast spindle density, and memory performance, distinguished Clusters 2 and 3 from Cluster 1, spindle amplitude was similar across Clusters 1 and 2. However, in Cluster 3, amplitude was markedly increased. As Cluster 3 was associated with worse nocturnal hypoxemia, we speculate that changes in spindle amplitude might be a prominent feature in people with greater sleep apnea severity. In our study, oxygen desaturation index was calculated as the number of times per hour blood oxygen levels dropped by 3% or more from baseline. Previous studies have reported similar findings with related sleep apnea severity measures. A higher degree of sleep apnea severity, as measured by the apnea-hypopnea index, has been associated with lower spindle density and greater spindle amplitude (40,42) while reduced spindle duration has also been associated with increasing sleep apnea severity (41). Reduced spindle amplitude is a biomarker of cognitive impairment and ageing, however it is less sensitive than spindle duration and fast spindle density (7,72). Sleep apnea is strongly associated with cognitive deficits (40,76) with nocturnal hypoxemia closely linked to impaired memory performance (77). While sleep apnea and cognitive decline may co-occur as part of an underlying neurodegenerative illness (78,79), cognitive deficits may also be secondary to or aggravated by sleep apnea (76,80,81). This picture fits with the Cluster 3 we identified, which included people with greater sleep apnea severity and reduced memory performance (although not as severe as Cluster 2). We speculate that higher spindle amplitude may be a feature in older people with an increased severity of sleep apnea, distinguishing them from people with a more significant, primary memory deficit.

Sleep quality, as measured by WASO, did not emerge as a significant predictor of cluster membership, although Figure 2a indicates that people with the highest WASO scores have higher probability of being in Cluster 2. This is supported by cluster group level differences (Table 1) confirming higher WASO scores in Cluster 2 relative to Cluster 3. Indeed, other markers of sleep quality detailed in Supplementary Material emphasise poor sleep quality in Cluster 2. In line with the hypothesis that sleep spindles protect sleep against environmental disturbances by elevating the arousal threshold, abnormalities in spindle achitecture may contribute to poor sleep quality (11,20,82).

Our resting state fMRI analysis revealed that default mode connectivity was related to spindle architecture, although this relationship differed across the cluster groups. Increased default mode connectivity was associated with fewer fast spindles and shorter spindle duration in Cluster 1 participants. However, for Clusters 2 and 3 increased default mode connectivity was associated with increased spindle duration, and in the case of Cluster 2, higher fast spindle density as well. We hypothesise that the different directional relationships captured here may reflect diverse underlying mechanisms, by which generally more intact spindles, seen in Cluster 1, may support more efficient default mode network function. These findings are in keeping with the mixed picture that emerges across default network resting state fMRI and clinical phenotypes in ageing and sleep. Both increased and decreased default network connectivity has been associated with impaired cognitive function in ageing (83,84) and with sleep disturbances (85–87). Neurodegenerative diseases of ageing present with a complex picture of both decreased and compensatory patterns of neuronal activation (88). These divergent patterns may drive distinct relationships across scales of measurement, i.e., across clinical measures, fMRI, and EEG, which may contribute to the variability seen in ageing and sleep disorders. As a key site of both structural and functional changes across neurodegenerative diseases of ageing (89,90), continued understanding of the bidirectional influence between default mode connectivity and sleep microarchitecture, may improve the capacity for these measures to complement each other as part of early diagnostic and monitoring strategies. Our findings highlight the need to account for different clinical phenotypes when assessing the relationship between sleep microarchitecture and fMRI connectivity in older adults.

Spindles occur across complex spatiotemporal scales, extending throughout multiple brain regions. Here we used central EEG recordings of spindles above the medial temporal lobe (C3 and C4) in an attempt to best measure fast spindle activity (4). Fast and slow spindles have distinct topographical distributions, with fast spindles showing a more centroparietal expression (15,16,91,92). Future studies should extend this investigation to other sites including frontal regions, which are more closely associated with slow spindles (4,92). Identifying variability in the topographical distribution and spatiotemporal patterns of both spindle types, and their coupling with slow oscillations, may better inform our understanding of the role of spindles in memory consolidation and perhaps glymphatic processes (93,94).

This study identified patterns in spindle architecture which are associated with distinct clinical phenotypes in older adults. We identified differences in default mode network connectivity across groups that inform the variability currently observed in the literature. Together, these findings link clinical measures, neuroimaging and sleep spindle architecture to distinct clinical phenotypes, enhancing our understanding of the bidirectional relationships between altered sleep and neurodegenerative diseases of ageing. Advances in this area will provide the impetus for future therapies that may target sleep spindles and/or nocturnal hypoxemia, with the possible utility for improving sleep as well as memory.

## Data Availability

All data produced in the present study are available upon reasonable request to the authors.

## Acknowledgements

This study was supported by the National Health and Medical Research Council (NHMRC) Centre of Research Excellence to Optimise Sleep in Brain Ageing and Neurodegeneration (CogSleepCRE GNT1152945). CO, AD and SLN were supported by NHMRC fellowships (2016866, 2008001, 2008064 respectively). The authors acknowledge the statistical assistance provided by the Sydney Informatics Hub, a Core Research Facility of the University of Sydney.

