## Supplementary Material for "Sleep spindle architecture associated with distinct clinical phenotypes in older adults at risk for dementia"

**1.** Resting state fMRI pre-processing	Page 2

**2.** Cluster identification consensus-based algorithm results	Page 3

**3.** Additional neuropsychological and sleep micro- and macro - architecture measures	Page 4

**4.** Resting state fMRI supplementary analysis	Page 5

**1. Resting state fMRI pre-processing**

Pre-processing was performed in MATLAB using the functional connectivity toolbox (CONN v19.c; <http://www.nitrc.org/projects/conn>).

Pre-processing involved a standardised Montreal Neurological Institute (MNI)-space direct normalisation pipeline (described in detail here (69). Briefly, each volume was co-registered and resampled to a reference volume (i.e., the first scan) using b-spline interpolation. Rigid head movement timeseries were calculated (6 degree of freedom) and added as a first-level covariate for motion correction (see aCompCor below). Slice-timing correction using sinc-interpolation to match the mid-TR time was then performed. Outlier scans were identified and defined as volumes with frame-wise displacement greater than 0.5mm or signal intensity changes greater than three standard deviations. Functional and structural data were the normalised to MNI standard space and segmented into grey matter, white matter, and cerebrospinal fluid using the SPM12 unified segmentation and normalisation procedure, which includes estimating the best non-linear spatial transformation (70). This procedure was applied to the functional data using the mean blood-oxygen-level-dependent signal as the reference image, and to the structural data using the raw T1-weighted volume as the reference image. Data were resampled to 2 mm isotropic voxels for functional data and 1 mm isotropic voxels for structural data, using 4th order spline interpolation.

A standard denoising pipeline was applied, involving two steps. First, aCompCor was applied to the BOLD timeseries to regress out noise and motion artefacts. Noise regressors were obtained from white matter and cerebrospinal fluid timeseries, as well as the first-level covariates previously defined (12 components from the estimated subject- motion parameters, derived from three translation and three rotation parameters and their first- order derivatives, and the outlier volumes to be ‘scrubbed’). Second, a temporal band pass filter was applied to the BOLD signal (0.009 – 0.08 Hz) after confound regression to minimise the influence of physiological, head-motion and other noise sources. This was performed after regression to avoid any frequency mismatch in the nuisance regression procedure (71). Images were then visually inspected for quality. Four participants were excluded from further analysis due to poor quality data or artefacts.

**2. Cluster identification consensus-based algorithm results**

An optimal solution of 3 clusters was identified by the consensus-based algorithm. The choice of 3 clusters is supported by 10 (34.48%) methods out of 29 (Ch, Hartigan, Scott, trcovw, Tracew, Ratkowsky, Ball, SDindex, Mixture (VVI), Mixture (EVI)).

**Supplementary Figure 1**

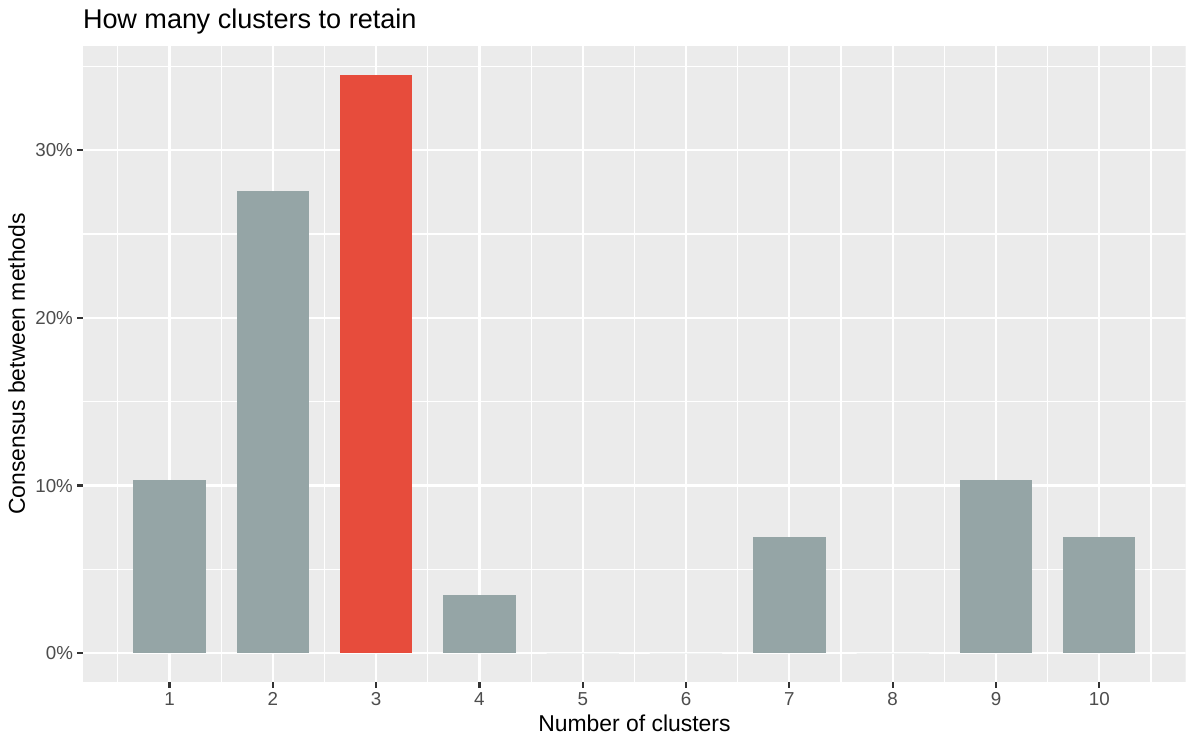

***Optimal number of clusters: method agreement procedure****.*

**3. Additional neuropsychological and sleep macro- and micro-architecture measures**

**Supplementary Table 1**

|  |  | **Group 1** | **Group 2** | **Group 3** | ***p* value** | **Pairwise** |
| --- | --- | --- | --- | --- | --- | --- |
| Learning | Logical Memory I, raw score | 42.07 (8.89) | 30.47 (12.44) | 33.22 (13.18) | **< .001** | *1 vs 2; 1 vs 3* |
|  | RAVLT 1-5, raw score | 51.29 (8.69) | 39.23 (12.73) | 43.61 (13.09) | **< .001** | *1 vs 2* |
| Executive function | FAS, total words | 42.41 (13.20) | 37.21 (9.99) | 44.11 (15.53) | .078 | - |
|  | Digit Span, total | 16.58 (3.81) | 16.46 (3.71) | 17.00 (4.77) | .890 | - |
|  | Trailmaking Part B, secs | 79.24 (30.56) | 99.82 (38.74) | 84.91 (33.40) | .051 | - |

***Additional neuropsychological characteristics of k-means defined cluster groups.*** *Data presented as mean (standard deviation). Group differences compared using one-way analysis of variance with post hoc Tukey’s honestly significant difference. P-values shown for group level comparisons, with significant (p < .05) pairwise comparisons noted. Learning was assessed via i) the sum of the five learning trials the Rey Auditory Verbal Learning Test (RAVLT 1-5; (1)); ii) the encoding part of the Logical Memory subtest of the Wechsler Memory Scale - III (2). Executive function was assessed via: i) Controlled Oral Word Association Test, phonemic fluency (FAS; (3)), with the total amount of words generated across three 1-minute periods the outcome of interest; ii) working memory maintenance and manipulation, as measured by score from the forwards and backwards digit span task (Digit Span; (2)); iii) attentional set shifting, measured by the Trail Making Test Part B (Trails B; (4)).*

**Supplementary Table 2**

| **Macroarchitecture** | **Cluster 1** | **Cluster 2** | **Cluster 3** | ***p* value** | **Pairwise** |
| --- | --- | --- | --- | --- | --- |
| Total time in bed (min) | 427.63 (51.14) | 461.46 (58.23) | 421.40 (46.29) | **0.010** | *1 vs 2; 2 vs 3* |
| Total sleep time (min) | 341.60 (54.7) | 346.3 (75.3) | 349.2 (55.9) | 0.920 | - |
| Stage N1 sleep (min) | 20.20 (16.77) | 24.85 (19.78) | 17.87 (10.62) | 0.300 | - |
| Stage N2 sleep (min) | 185.00 (42.67) | 161.90 (46.73) | 198.20 (57.36) | **0.018** | *2 vs 3* |
| Stage N3 sleep (min) | 75.98 (27.04) | 84.69 (48.08) | 76.78 (36.86) | 0.620 | - |
| REM sleep (min) | 60.40 (21.22) | 70.73 (30.32) | 52.55 (28.89) | 0.052 | *-* |
| Sleep efficiency (%) | 80.22 (11.54) | 74.80 (13.46) | 83.07 (11.03) | **0.044** | *2 vs 3* |
| Sleep latency (min) | 23.15 (25.72) | 23.37 (25.13) | 25.54 (28.87) | 0.950 | - |
| REM latency (min) | 136.60 (81.37) | 108.8 (63.17) | 137.90 (86.12) | 0.220 | - |
| AHI (events/hour) | 13.52 (13.86) | 14.65 (14.29) | 24.97 (19.94) | **0.041** | - |

***Sleep macroarchitecture characteristics of k-means defined cluster groups.*** *Data presented as mean (standard deviation). Group differences compared using one-way analysis of variance with post hoc Tukey’s honestly significant difference. P-values shown for group level comparisons, with significant (p < .05) pairwise comparisons noted. Sleep efficiency calculated as ratio of total sleep time to time in bed, multiplied by 100 to display as a percentage NREM min calculated as sum of N2 min and N3 min. AHI; Apnea hypopnea index calculated as the number of apneas and hypopneas per hour of total sleep time.*

**Supplementary Table 3**

| **Spindle architecture** | **Cluster 1** | **Cluster 2** | **Cluster 3** |
| --- | --- | --- | --- |
| NREM fast spindle density (events per min) | 1.10 (0.55) | 0.21 (0.26) | 0.30 (0.24) |
| NREM spindle amplitude (µV) | 15.87 (3.15) | 13.84 (3.25) | 24.81 (5.54) |
| NREM spindle duration (seconds) | 0.81 (0.03) | 0.72 (0.03) | 0.75 (0.04) |

***Sleep spindle architecture of k-means defined cluster groups.*** *Data presented as mean (standard deviation).*

**4. Resting state fMRI supplementary analysis and figures**

**Supplementary Table 4**

| **Spindle measure** | **Network** | **Main effect (connectivity)** | **Interaction effect (connectivity: cluster)** |
| --- | --- | --- | --- |
| Fast density | CO | F(1,76) = 0.36, *p* = 0.552 | F(1,76) = 0.40, *p* = 0.671 |
|  | DAN | F(1,76) = 0.13, *p* = 0.724 | F(1,76) = 0.02, *p* = 0.985 |
|  | FPN | F(1,76) = 0.48, *p* = 0.492 | F(1,76) = 0.06, *p* = 0.940 |
|  | RSP | F(1,76) = 0.90, *p* = 0.347 | F(1,76) = 1.33, *p* = 0.271 |
|  | VAN | F(1,76) = 1.64, *p* = 0.204 | F(1,76) = 0.78, *p* = 0.463 |
| Duration | CO | F(1,76) = 2.47, *p* = 0.120 | F(1,76) = 1.15, *p* = 0.323 |
|  | DAN | F(1,76) = 3.06, *p* = 0.084 | F(1,76) = 0.88, *p* = 0.419 |
|  | FPN | F(1,76) = 0.02, *p* = 0.898 | F(1,76) = 0.56, *p* = 0.574 |
|  | RSP | F(1,76) = 2.04, *p* = 0.158 | F(1,76) = 1.30, *p* = 0.279 |
|  | VAN | F(1,76) = 0.85, *p* = 0.359 | F(1,76) = 1.60, *p* = 0.208 |

***Output from additional functional connectivity linear models.*** *Models examining main effect of functional connectivity and interaction effect of functional connectivity and cluster membership on sleep spindle characteristics. Amplitude not investigated as no significant effect of default mode connectivity emerged. CO = cingulo opercular, DMN = dorsal mode network, DAN = dorsal attention network, FPN = frontoparietal network, RSP = Retrosplenial network VAN = ventral attention network. Salience network excluded from further analysis due to small size (number of regions = 5).*
